# Cognitive impairment at older ages among 8000 men and women living in Mexico City: cross-sectional analyses of a prospective study

**DOI:** 10.1101/2024.04.25.24306237

**Authors:** Carlos González-Carballo, Pablo Kuri-Morales, Erwin Chiquete, Mario Rojas-Russell, Rogelio Santacruz-Benitez, Raúl Ramirez-Reyes, Adrián Garcilazo-Ávila, Jaime Berumen, Eirini Trichia, Louisa Gnatiuc Friedrichs, Paulina Orellana, Carolina Ochoa-Rosales, Gary O’Donovan, Jonathan R Emberson, Roberto Tapia-Conyer, Diego Aguilar-Ramirez, Jesus Alegre-Díaz

**Affiliations:** Experimental Research Unit from the Faculty of Medicine, National Autonomous University of Mexico, Mexico City, Mexico; Instituto Tecnológico y de Estudios Superiores de Monterrey, Monterrey, México; Instituto Nacional de Ciencias Médicas y Nutrición Salvador Zubirán, México City, México; Faculty of Higher Studies Zaragoza, National Autonomous University of México, México City, México; Clinical Trial Service Unit and Epidemiological Studies Unit, Nuffield Department of Population Health, University of Oxford, Oxford, UK; Latin American Brain Health Institute (BrainLat), Universidad Adolfo Ibanez, Santiago, Chile; Center for Social and Cognitive Neuroscience (CSCN), School of Psychology, Universidad Adolfo Ibanez, Santiago, Chile; Facultad de Medicina, Universidad de los Andes, Bogotá, Colombia

## Abstract

**Importance:** There is limited population-based evidence on the prevalence of cognitive impairment in Mexico, a country with a rapidly aging population and where key risk factors, such as diabetes and obesity, are common.

**Objective:** To describe the distribution of cognitive impairment in a sample of adults from Mexico City.

**Design, Setting, and Participants:** This cross-sectional population-based study included participants from the Mexico City Prospective Study which 50,000 men and 100,000 women aged ≥35 years from two districts in 1998-2004. In 2015-2019 about 10,000 survivors were resurveyed with identical information from the original survey and additional assessments including a cognitive assessment. The main analyses included those aged 50-89 years with complete cognitive assessment and covariate data at resurvey.

**Main outcomes and measures:** Cognition was assessed using the Mini Mental State Examination (MMSE) and those with cognitive impairment (MMSE ≤24) were identified. The distribution of MMSE scores and cognitive impairment by age, sex, and major disease risk factors (diabetes, hypertension, and adiposity) was analyzed among those with complete MMSE data and some degree of self-reported formal education.

**Results:** Of the 9,288 participants aged 50-89 years at the 2015-2019 resurvey with complete data, 8,197 reported having at least some years of formal education. Among these (mean age 66 years; 31% men), their mean MMSE score was 26.2 (SD 3.6) points, 1,941 (24%) had cognitive impairment, mean body-mass index (BMI) was 28.6 (SD 5.5) kg/m^2^, 3,008 (37%) had previously-diagnosed hypertension and 2,467 (30%) had previously-diagnosed diabetes. The sex- and district-standardised prevalence of cognitive impairment increased strongly with age, from 10% in those 50-59 years to 55% in those aged 80-89. At any given age, the prevalence of cognitive impairment was higher in women than in men. After accounting for the effects of age, sex, and district there was little difference in the prevalence of cognitive impairment between participants with or without diabetes, hypertension, overweight or obesity (BMI ≥25 km/m^2^), or high levels of fat mass.

**Conclusion and relevance:** In this population of adults aged 50-89 years from two districts of Mexico City, the prevalence of cognitive impairment was high, particularly among women. The extent to which cognitive impairment relates to health outcomes in this population needs to be investigated.

**KEY POINTS:** *Question:* What is the distribution of cognitive impairment among adults in Mexico City?

*Findings:* In this cross-sectional study of 8,197 adults aged 50-89 years, the sex- and district-standardised prevalence of cognitive impairment ranged from 10% in those 50-59 years to 55% in those aged 80-89 and at any given age was higher in women than in men. There was little difference in the prevalence of cognitive impairment between participants with or without hypertension, diabetes or excess of adiposity.

*Meaning:* Cognitive impairment is common in adults in Mexico City. Its relevance to major morbidity and mortality deserves future research.

## INTRODUCTION

Epidemiological and demographic changes in global populations have highlighted the relevance of the ageing process for the preservation of cognitive function, which has a significant effect on the independence and quality of life of older people ^1–3^. Metabolic diseases, such as excess adiposity or type 2 diabetes, and cardiovascular diseases have been identified as risk factors for the development of cognitive impairment ^4–6^, a condition which has been associated with an increased risk of dementia ^7,8^. Given how rapidly populations across the world are aging, including those in understudied regions such as Latin America, there is a need for a greater understanding of the distribution of cognitive impairment and its associated risk factors in such regions.

Mexico is the second most populous country in Latin America at around 126 million people.^9^ Obesity and diabetes are common in Mexico and confer an unduly high excess risk of mortality (mostly due to vascular and renal causes)^10–12^. Obesity^13,14^ and diabetes^15^ are also established risk factors for cognitive impairment and dementia. The reported prevalence of cognitive impairment in Mexico is estimated to be around 7% to 10% in those aged 65 years increasing to 27% in those aged ≥85 years ^16,17^. However, these estimates are based on studies with very small sample sizes^17^ or where information on cognition was not exclusively collected directly from participants but also through proxy interviews^16^.

The Mexico City Prospective Study (MCPS) recruited 150,000 participants in 1998-2004. We aimed to report the age- and sex-specific distribution of cognitive impairment and its sociodemographic, lifestyle, biological and medical health characteristics in a subset of around 9,000 MCPS participants who had cognitive function assessed during a resurvey of survivors carried out in 2015-2019.

## METHODS

### Study design and participants

Study design, sampling methods, and follow-up have been described before^18^. Briefly, between 1998 and 2004, households in two contiguous districts in Mexico City (Coyocán and Iztapalapa) were visited and all inhabitants aged 35 and over were invited to participate in the study. Of the 112,333 households visited, 106,059 (94%) yielded at least one inhabitant who consented to participate. Ethics approval was granted by the Mexican Ministry of Health, the Mexican National Council for Science and Technology, and the University of Oxford. All participants provided written informed consent.

### Data collection

In the baseline measurement, during household visits, trained nurses conducted face-to-face interviews and recorded information on sociodemographic and lifestyle factors, current medications, and medical history (including previous medically diagnosed diseases such as diabetes). Blood pressure, weight, height, waist and hip circumference were measured using standard protocols, and a non-fasting venous 10 mL blood sample was collected. In 2015-2019, a repeat survey was performed in a subset of around 10,000 surviving participants. For this resurvey, streets within the two study districts in which participants were previously recruited in 1998-2004 were selected at random and then systematically revisited to identify participants who were still alive, living at the same address, and willing to take part in a further survey. Data from the resurvey were entered directly into study tablets which captured the exact time points at which each question or procedure was completed. The resurvey involved similar questions and assessments as at the original survey in 1998-2004, but with some additions. Whole-body bio-impedance measures at resurvey were recorded using a Tanita SC240-MA body composition analyzer. Cognition at resurvey was assessed using the Mini Mental State Examination (MMSE) in Latin-American Spanish^19^, a tool that is commonly used to identify individuals at a higher risk of cognitive impairment in clinical and population studies. The original MMSE^20^ and the version used in this study have the same domains and scores: orientation to time (5 points); orientation to place (5 points); registration (3 points); attention and calculation (5 points); recall (3 points); and, language (9 points). Cognitive impairment was defined as a score of 24 or less on the MMSE, to be consistent with previous studies in Mexican population^19,21^.

### Statistical analysis

Analyses were limited to resurveyed individuals whose cognitive status was assessed with the MMSE. The few participants who were aged below 50 years or were aged 90 years or above were excluded. We also excluded anyone with an MMSE score of zero, anyone with an MMSE questionnaire duration less than 1 minute or longer than 30 minutes, and anyone with a missing self-reported education (**Supplementary Table 1**).

Initial analyses plotted the distributions of MMSE score and of cognitive impairment at different ages across groups of educational attainment reported at resurvey (i.e., without formal education, incomplete primary education, complete primary education, secondary education, and tertiary education). As having no formal education strongly limited the ability of participants to complete the MMSE questionnaire reliably (**Supplementary Figure 1**), the main analyses were limited to the large proportion of participants with some degree of formal education. (Analyses including those reporting no formal education are provided as sensitivity analyses.)

The prevalence of cognitive impairment and the mean MMSE scores (with 95% confidence intervals) were plotted separately in men and women across equally sized age groups (∼1000 women and ∼500 men in each group). To obtain standardised estimates, the prevalence of cognitive impairment was first calculated separately in men and women at ages 50-59, 60-69, 70-79, and 80-89 years. These eight age- and sex-specific estimates were obtained by averaging the group-specific prevalences in participants residing in Coyoacán and in Iztapalapa, yielding district-standardised estimates. These were then averaged to calculate the overall age-, sex- and district-uniformly-standardised prevalence at ages 50-89 years.

Finally, to assess how the prevalence of cognitive impairment varied in those with and without key risk factors, while accounting for variation due to age, sex, and district of residence, sex- and district-standardised prevalences across equally-sized age groups were calculated in those with and without such factors. The factors included previously-diagnosed diabetes (i.e., a self-reported medical diagnosis or treatment of diabetes), previously-diagnosed hypertension (i.e., a self-reported medical diagnosis or treatment of hypertension), overweight or obesity (i.e., a body-mass index of 25 kg/m^2^ or above) and high impedance-measured fat mass (i.e., those at the top half of the distribution of fat mass). Analyses were conducted using R (version 4.3.2).

## RESULTS

Of the 159,755 participants recruited at baseline from 1998 to 2005, 10,144 were resurveyed in 2015-2019. Of these, 388 participants were aged 49 years or below or 90 years or above at resurvey, 218 had missing MMSE score, 370 had a very short or very long MMSE questionnaire duration, and 126 had missing data on education, and were excluded. Of the remaining 9,288 men and women aged 50 to 89 years, 1,091 reported not having had any formal education and were removed (**Supplementary Figure 1**), leaving 8,197 participants for the main analyses.

**Table 1** shows the characteristics at resurvey of the 2,565 men and 5,632 women aged 50 to 89 years who reported having some level of formal education. In these participants, the mean (SD) MMSE score was 26.2 (3.6) in both men and women (**Supplementary Figure 2**), the mean (SD) age was 66 (9.7) years, about half were residents of Coyoacán, and 11% had completed tertiary-level education. About one in six (14%) were current smokers, nearly half (44%) were current drinkers, and 17% engaged in leisure-time physical activity at least 3 times per week. In the study population, obesity, hypertension, and diabetes were common. The mean (SD) body-mass index (BMI) was 28.6 (5.5) kg/m^2^, about half of the population (42%) had BMI between 25 and 29.9 kg/m^2^, and one-third (35%) had a BMI ≥30 kg/m^2^; the mean (SD) systolic blood pressure (SBP) was 132 (22) mmHg and about a third (30%) had previously diagnosed diabetes. Compared with women, men were more likely to have tertiary-level education (13% vs 9%) to smoke (24% vs 10%), and to drink alcohol (56% vs 38%). However, women, on average, had higher BMI (29.2 vs 27.5 kg/m^2^) and were more likely to have previously diagnosed hypertension (39% vs 32%) than men.

**Table 1.** Characteristics at resurvey of 8,197 men and women aged 50-89 with MMSE information and some level of education, by cognitive impairment status (MMSE ≤24)

|  | Men |  |  | Women |  |  | Overall |  |  |
| --- | --- | --- | --- | --- | --- | --- | --- | --- | --- |
| | With cognitive impairment<br>MMSE ≤24<br>(N=588) | Without cognitive impairment<br>MMSE >24<br>(N=1977) | All men<br>(N=2565) | With cognitive impairment<br>MMSE ≤24<br>(N=1353) | Without cognitive impairment<br>MMSE >24<br>(N=4279) | All women<br>(N=5632) | With cognitive impairment<br>MMSE ≤24<br>(N=1941) | Without cognitive impairment<br>MMSE >24<br>(N=6256) | All participants<br>(N=8197) |
| **MMSE, score** | 20.8 (3.4) | 27.8 (1.6) | 26.2 (3.6) | 21.1 (3.2) | 27.9 (1.6) | 26.2 (3.6) | 21.0 (3.2) | 27.9 (1.6) | 26.2 (3.6) |
| **Age, years** | 74 (9.4) | 65 (9.3) | 67 (9.9) | 71 (9.9) | 63 (8.7) | 65 (9.5) | 72 (9.8) | 64 (8.9) | 66 (9.7) |
| **Socio-economic status and lifestyle behaviors** |  |  |  |  |  |  |  |  |  |
| Resident of Coyoacán | 368 (63%) | 1049 (53%) | 1417 (55%) | 766 (57%) | 2065 (48%) | 2831 (50%) | 1134 (58%) | 3114 (50%) | 4248 (52%) |
| Education |  |  |  |  |  |  |  |  |  |
| Unfinished primary | 277 (47%) | 282 (14%) | 559 (22%) | 766 (57%) | 754 (18%) | 1520 (27%) | 1043 (54%) | 1036 (17%) | 2079 (25%) |
| Completed primary | 230 (39%) | 691 (35%) | 921 (36%) | 447 (33%) | 1702 (40%) | 2149 (38%) | 677 (35%) | 2393 (38%) | 3070 (37%) |
| Completed secondary | 67 (11%) | 676 (34%) | 743 (29%) | 106 (8%) | 1332 (31%) | 1438 (26%) | 173 (9%) | 2008 (32%) | 2181 (27%) |
| Completed tertiary | 14 (2%) | 328 (17%) | 342 (13%) | 34 (3%) | 491 (11%) | 525 (9%) | 48 (2%) | 819 (13%) | 867 (11%) |
| Widowed | 126 (21%) | 208 (11%) | 334 (13%) | 490 (36%) | 896 (21%) | 1386 (25%) | 616 (32%) | 1104 (18%) | 1720 (21%) |
| Current smoker | 117 (20%) | 509 (26%) | 626 (24%) | 88 (7%) | 465 (11%) | 553 (10%) | 205 (11%) | 974 (16%) | 1179 (14%) |
| Current drinker | 241 (41%) | 1182 (60%) | 1423 (56%) | 404 (30%) | 1761 (41%) | 2165 (38%) | 645 (33%) | 2943 (47%) | 3588 (44%) |
| Leisure-time physical activity, ≥3 times per week | 93 (16%) | 406 (21%) | 499 (20%) | 139 (10%) | 716 (17%) | 855 (15%) | 232 (12%) | 1122 (18%) | 1354 (17%) |
| **Physical measurements\*** |  |  |  |  |  |  |  |  |  |
| Body-mass Index, kg/m<sup>2</sup> | 26.9 (5.0) | 27.7 (5.0) | 27.5 (5.0) | 28.4 (5.9) | 29.4 (5.6) | 29.2 (5.7) | 28.0 (5.7) | 28.8 (5.5) | 28.6 (5.5) |
| BMI 25-29.9 | 206 (46%) | 772 (49%) | 978 (48%) | 395 (38%) | 1399 (40%) | 1794 (39%) | 601 (40%) | 2171 (43%) | 2772 (42%) |
| BMI ≥30 | 92 (20%) | 402 (26%) | 494 (24%) | 366 (35%) | 1444 (41%) | 1810 (40%) | 458 (31%) | 1846 (36%) | 2304 (35%) |
| Waist-to-hip ratio, cm | 0.99 (0.083) | 0.98 (0.057) | 0.98 (0.064) | 0.94 (0.074) | 0.92 (0.068) | 0.93 (0.070) | 0.95 (0.079) | 0.94 (0.069) | 0.94 (0.072) |
| Fat mass, kg | 18.5 (8.2) | 20.5 (8.5) | 20.1 (8.4) | 23.1 (9.7) | 25.8 (9.5) | 25.2 (9.6) | 21.7 (9.5) | 24.2 (9.5) | 23.6 (9.5) |
| Muscle mass, kg | 48.0 (7.1) | 50.6 (6.9) | 50.0 (7.1) | 37.3 (4.8) | 38.9 (4.6) | 38.5 (4.7) | 40.5 (7.4) | 42.5 (7.7) | 42.1 (7.7) |
| **Blood pressure** |  |  |  |  |  |  |  |  |  |
| SBP, mmHg | 135.8 (23.8) | 132.5 (20.7) | 133.2 (21.5) | 136.9 (24.2) | 130.1 (21.0) | 131.7 (22.0) | 136.5 (24.0) | 130.9 (20.9) | 132.2 (21.8) |
| DBP, mmHg | 78.4 (11.8) | 80.9 (11.0) | 80.3 (11.3) | 79.0 (11.3) | 79.8 (10.6) | 79.6 (10.8) | 78.8 (11.4) | 80.1 (10.7) | 79.8 (10.9) |
| Previously diagnosed or treated hypertension | 217 (37%) | 593 (30%) | 810 (32%) | 613 (45%) | 1585 (37%) | 2198 (39%) | 830 (43%) | 2178 (35%) | 3008 (37%) |
| **Diabetes** |  |  |  |  |  |  |  |  |  |
| HbA1c, % | 5.7 [5.4-6.5] | 5.6 [5.4-6.4] | 5.7 [5.4-6.4] | 5.8 [5.4-6.6] | 5.7 [5.4-6.4] | 5.7 [5.4-6.5] | 5.8 [5.4-6.6] | 5.7 [5.4-6.4] | 5.7 [5.4-6.5] |
| Previously diagnosed or treated diabetes | 193 (33%) | 544 (28%) | 737 (29%) | 456 (34%) | 1274 (30%) | 1730 (31%) | 649 (33%) | 1818 (29%) | 2467 (30%) |

MMSE, Mini-Mental State Examination; BMI, body mass index; SBP, systolic blood pressure; DBP, diastolic blood pressure; SD, standard deviation; IQR, interquartile range. Numbers shown are *n* (%), mean (SD) or median (IQR). Exclusion criteria described in Supplementary Table 1. Missing data percentages were: BMI (20%), waist-hip-ratio (3%), fat mass (19%), muscle mass (19%), and blood pressure (1%).

Of all 8,197 participants included in the main analyses, 1,941 (23%) had an MMSE score ≤24 consistent with cognitive impairment. Those with cognitive impairment were, on average, about 8 years older than those without (72 [9.8] vs 64 [8.9]), were more likely to be residents of Coyoacán (58% vs 50%) and had lower educational attainment (2% vs 13% with tertiary-level education). They were also more likely to be widowed (32% vs 18%), and less likely to smoke (11% vs 16%), drink (33% vs 47%), or exercise (12% vs 18% had leisure-time physical activity ≥3 times per week). Participants with cognitive impairment also had a lower BMI (28.0 vs 28.8 kg/m^2^), were less likely to have obesity (31% vs 36%) and were more likely to have previously diagnosed hypertension (43% vs 35%) and previously diagnosed diabetes (33% vs 29%).

### Age and sex specific prevalence of cognitive impairment

The overall age-, sex-, and district-standardised prevalence of cognitive impairment at ages 50-89 years was 29% and was moderately higher in women (31%) than in men (26%) (**Table 2**). In both men and women, the prevalence increased steeply with age, from 8% at ages 50-59 years to 52% at ages 80-89 years in men and, just as steeply, from 12% at ages 50-59 years to 59% at ages 80-89 years in women (**Table 2 and Figure 1**). When averaged across men and women, the mean MMSE score reduced from 27.6 (2.3) at ages 50-59 years to 23.0 (4.8) at ages 80-89 years.

**Figure 1.**
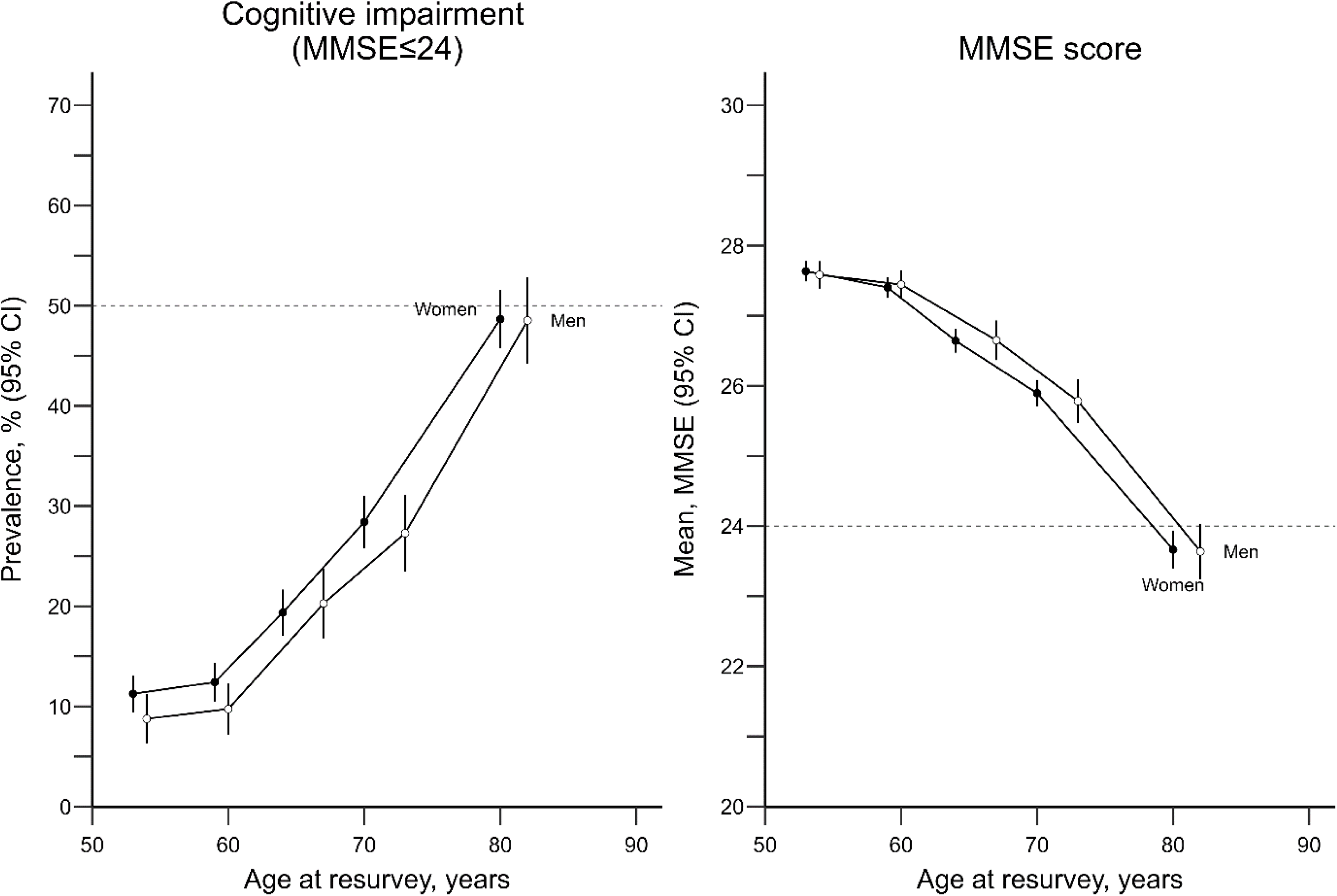
District-standardised sex-specific prevalences of cognitive impairment and mean MMSE score in 8,197 participants aged 50 to 89 years who reported having had some level of formal education. MMSE, Mini-Mental State Examination; CI, confidence interval. Each estimate is the average between participants form Coyoacan and Iztapalapa, yielding district-standardised age- and sex-specific estimates. Exclusions and conventions as per Table 1. Each point involves ∼1050 women and ∼470 men.

**Table 2.** Age- and sex-specific mean MMSE score and prevalences of cognitive impairment in 8,197 participants aged 50 to 89 years at resurvey who reported having had at least some level of formal education

| Age group | Men |  |  | Women |  |  | Overall |  |  |
| --- | --- | --- | --- | --- | --- | --- | --- | --- | --- |
|  | n | MMSE |  | n | MMSE |  | N | MMSE |  |
|  |  | mean | Prevalence |  | mean | Prevalence |  | mean | Prevalence |
|  |  | (SD) | % |  | (SD) | % |  | (SD) | % |
| 50-59 | 695 | 27.6 (2.2) | 8 | 1828 | 27.5 (2.5) | 12 | 2523 | 27.6 (2.4) | 10 |
| 60-69 | 804 | 27 (2.9) | 16 | 1990 | 26.7 (2.9) | 19 | 2794 | 26.9 (2.9) | 18 |
| 70-79 | 724 | 25.5 (3.8) | 30 | 1286 | 25.2 (3.7) | 34 | 2010 | 25.3 (3.8) | 32 |
| 80-89 | 342 | 23.3 (4.6) | 52 | 528 | 22.6 (5) | 59 | 870 | 23.0 (4.8) | 55 |
| **All** | **2565** | **25.9 (3.4)** | **26** | **5632** | **25.5 (3.5)** | **31** | **8197** | **25.7 (3.5)** | **29** |

MMSE, Mini-Mental State Examination. Cognitive impairment is defined as MMSE score $\leq 24$ .
Age- and sex-specific estimates are the simple average of such estimates in each district, giving district-standardised estimates. For men and for women, the four district-standardised age-specific estimates are subsequently averaged to give age- and district-standardised sex-specific estimates. Then, the eight district-standardised age- and sex-specific estimates are averaged to give an overall uniformly-standardised estimate.

### Cognitive impairment by key metabolic risk factors

After standardising for age, sex, and district of residence, there was little to no difference in the prevalence of cognitive impairment in individuals with key metabolic risk factors compared with those without (**Figure 2**). At every given age, the sex- and district-standardised prevalence of cognitive status was broadly similar in individuals with and without previously diagnosed hypertension, with and without a BMI ≥25 kg/m^2^, and with and without a fat mass above the sex-specific median (>19.2 kg in men or >24.4 kg in women). There was some suggestion that the prevalence of cognitive impairment between ages 60 and 75 was slightly higher in those with versus without previously-diagnosed diabetes, but such differences were not seen at either younger or older ages.

**Figure 2.**
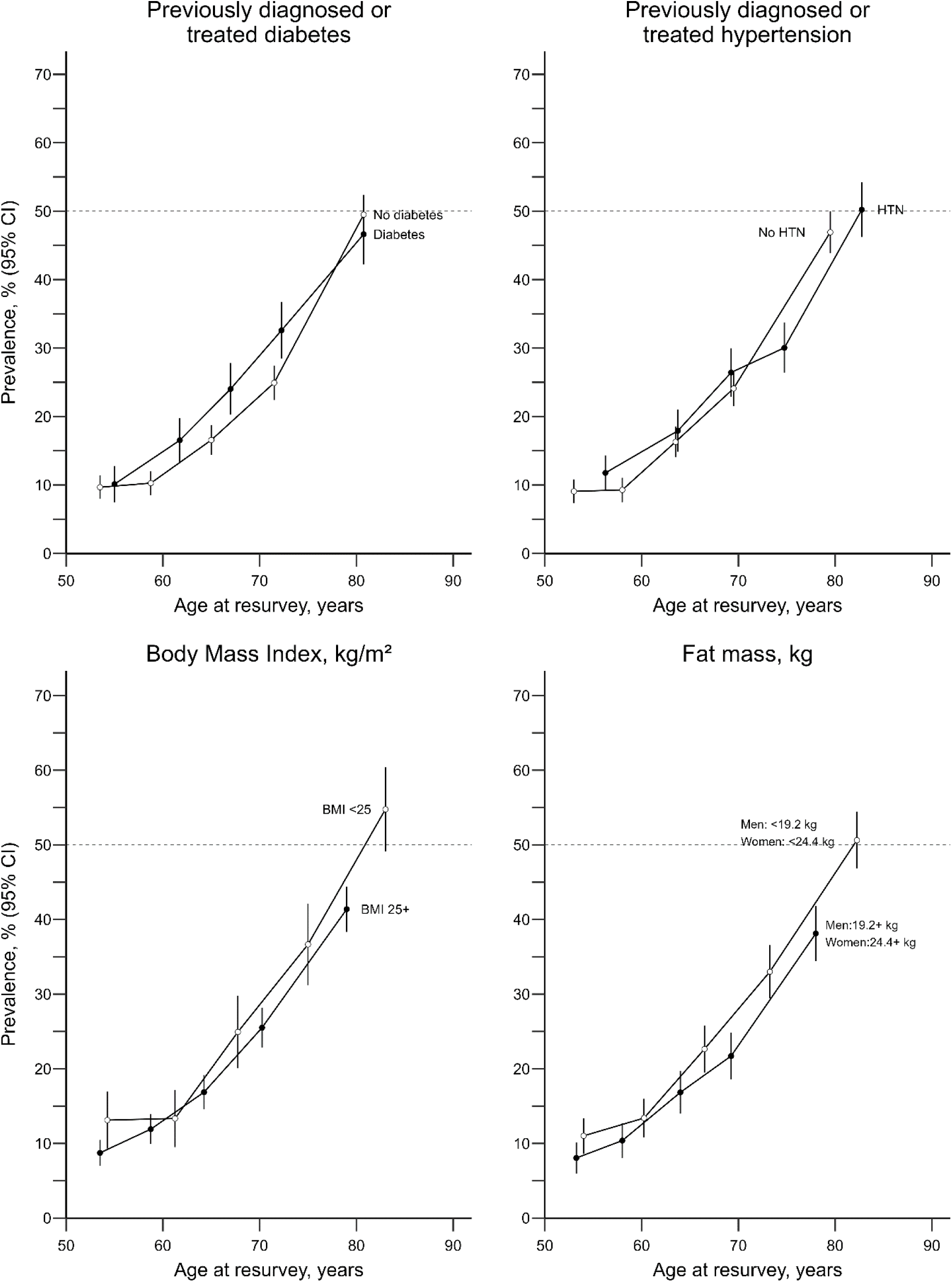
Sex- and district-standardised prevalence of cognitive impairment in 7,628 participants aged 50 to 89 years at resurvey who reported having had some level of education with and without key metabolic risk factors, by age. MMSE, Mini-Mental State Examination; CI, confidence interval; HTN, hypertension. Sex-specific median fat mass (19.1 kg for men and 24.4 kg for women) were used as cut-offs. Estimates shown among 8,197 participants for the top-row panels, 6,586 participants for the bottom-left panel, and 6,676 for the bottom-right panel.

### Sensitivity Analyses

The prevalence of cognitive impairment showed similar distributions by age and sex in residents from the two districts, but the overall age- and sex-standardised prevalence was higher in Coyoacán (32%) than in Iztapalapa (25%) (**Supplementary Table 2**). When considering all the population with and without formal education, the overall sex- and district-standardised prevalence increased to 35% (31% in men and 39% in women; **Supplementary Table 3**).

## DISCUSSION

This study describes the distribution of cognitive impairment among 9000 participants of the Mexico City Prospective Study whose cognition was assessed with the Mini-Mental State Examination in 2015-2019. The uniformly-standardised prevalence of cognitive impairment at age 50-89 years was 29% overall, increasing from 10% at ages 50-59 years to 55% at ages 80-89 years, and was higher in women than in men overall (31% vs 26%, respectively) and at every given age. After accounting for age, sex and district of residence, the prevalence of cognitive impairment was broadly similar in those with vs without previously diagnosed diabetes, hypertension, BMI ≥25 kg/m^2^, or an above-average fat mass. Overall, the findings from this study highlight that, in this urban Mexican population, cognitive impairment is common.

This study builds on previous studies of cognitive impairment in Mexico. While it is difficult to compare directly, other studies in the Mexican population have reported a wide range of prevalences of cognitive impairment in adults (7%-34%). Differences across studies may be explained by different study characteristics (particularly differences in age which may not have been accounted for through standardization), different tools used for identifying cognitive impairment, and, in some cases, limited study sample size (which can yield unreliable estimates) ^5,22,23^. Our results are similar to those reported by a study that used data from the Mexican Health and Aging Study (MHAS), which included 10,000 adults aged 60 years or above from different regions in Mexico. In this study ^24^, cognition was evaluated in 2015 with the Cross-Cultural Cognitive Evaluation questionnaire (CCCE) through either direct or proxy interviews. The prevalence of any cognitive alteration (i.e.,cognitive impairment or dementia) was 20% and increased with age (17% at ages 60-75 years and 30% at ages >75 years). Another study that used data from a subsample of 1,800 MHAS participants assessed in 2015, reported a 34% prevalence of cognitive impairment in adults aged 60 years or above, perhaps because more comprehensive assessments were used to identify those with cognitive impairment ^6^. Pooled together, our study further suggest that cognitive impairment is common among adults in Mexico City and highlights the steep increase in its prevalence as individuals age.

Consistent with previous studies, we found a higher prevalence of cognitive impairment in women than men.^6,24^ There are many reasons why women may be more likely to develop cognitive impairment than men, particularly in low and middle-income countries. In the current study, individuals were born between ∼1925 and ∼1965. During this period, women in Mexico may have had lower opportunities to access higher levels of education than men, a strong risk factor for developing cognitive impairment. Women in the current study were slightly less likely than men to have tertiary-level education for instance. Women in Mexico may also be more likely to be unpaid for their labour, have poorer career progression opportunities and less likely to be economically independent than men^25^. Such factors have been associated with higher risk of cognitive impairment ^26^. While it is not fully understood how low education and socio-economic status lead to cognitive impairment, it is hypothesized that these factors limit the development of cognitive reserve in early and middle-life which could be protective of dementia ^27^. It is also likely that socio-economic status can increase the risk of cognitive impairment (and dementia) due to social disparities in smoking, alcohol intake, physical activity, adiposity, elevated blood pressure, and diabetes all of which are established risk factors for dementia ^28^. Overall, our findings highlight the potential benefits of addressing gender and socio-economic disparities to close health gaps.

After accounting for age, there was little difference in the prevalence of cognitive impairment in those with vs without hypertension or excess of adiposity. This may be expected if presence of these risk factors in early and middle-life is more relevant to cognitive impairment than presence in older age. This may be particularly relevant for adiposity, for which long-term prospective evidence has shown that those who develop dementia can lose weight many years before the diagnosis of the disease ^29^. Those with diabetes had a marginally higher prevalence of cognitive impairment than those without diabetes (at least between ages 60 to 75 years), but further research is needed to establish the extent to which diabetes may result in cognitive impairment in this population.

This study has several strengths, not least that it provides population-based prevalence estimates of cognitive impairment in an understudied population. Information was collected by trained health professionals using standardised procedures. While the tool used to assess cognition (MMSE) was developed in an English-speaking population from a high-income country, it has previously been validated for use in Spanish-speaking individuals ^19^. Although the full-length version of the MMSE was used, this tool substantially underestimates cognition level in individuals with little to no literacy skills (as certain tasks involve reading or writing). For this reason, we excluded from our main analyses participants who reported not having had any formal education. However, as a consequence of restricting our analyses to those who reported having at least some formal education, we may have even underestimated the true prevalence of cognitive impairment in the whole population. By design, the Mexico City Prospective Study is not representative of the whole population of Mexico. However, the standardised prevalence estimates, including age-, and sex-specific estimates, within this report facilitate comparison with other existing and future estimates and represent a valuable contribution to the epidemiological characterization of cognitive impairment in Mexico.

In conclusion, in this cross-sectional population-based study of Mexican adults aged 50-89 years, the prevalence of cognitive impairment assessed in 2015-2019 was high, particularly among women, and increased steeply with age. The extent to which cognitive impairment relates to health outcomes in this population needs to be investigated.

### Contributors

PK-M, JA-D, and RT-C established the cohort. PK-M, JRE, JA-D, and RT-C obtained funding. All authors contributed to data acquisition, analysis, or interpretation of data. CG-C and DA-R drafted the first version of the manuscript. All authors contributed to the critical revision of the manuscript for important intellectual content. All authors have seen and approved the final version and agreed to its publication. CG-C and DA-R had full access to all the data in the study, verified the underlying data, and take responsibility for the integrity of the data and the accuracy of the analysis.

### Disclosures

An unauthorised version of the Spanish MMSE was used by the MCPS study team without permission. The MMSE is a copyrighted instrument and may not be used or reproduced in whole or in part, in any form or language, or by any means without written permission of PAR.

### Data sharing

The Mexico City Prospective Study reflects a long-standing collaboration between researchers at the National Autonomous University of Mexico and the University of Oxford. Data from the Mexico City Prospective Study are available to bona fide researchers. The study’s Data and Sample Sharing policy can be downloaded (in English or Spanish: https://www.ctsu.ox.ac.uk/research/mcps). Available study data can be examined in detail through the study’s Data Showcase (https://datashare.ndph.ox.ac.uk/mexico/).

## Data Availability

The Mexico City Prospective Study reflects a long-standing collaboration between researchers at the National Autonomous University of Mexico and the University of Oxford. Data from the Mexico City Prospective Study are available to bona fide researchers. The study's Data and Sample Sharing policy can be downloaded (in English or Spanish: https://www.ctsu.ox.ac.uk/research/mcps). Available study data can be examined in detail through the study's Data Showcase. (https://datashare.ndph.ox.ac.uk/mexico/)

## Acknowledgments

We thank the study participants. The Mexico City Prospective Study has received funding from the Mexican Health Ministry, the National Council of Science and Technology for Mexico, the Wellcome Trust (058299/Z/99), Cancer Research UK, the British Heart Foundation (RE/13/1/30181), and the UK Medical Research Council (MC_UU_00017/2).

## Rights retention statement

For the purposes of open access, the authors have applied a Creative Commons Attribution (CC BY) licence to any Author Accepted Manuscript version arising.

