## Supplementary Material for "Cognitive impairment at older ages among 8000 men and women living in Mexico City: cross-sectional analyses of a prospective study"

### Supplementary Tables


1. Analysis inclusion criteria 2
2. Age- and sex-specific mean MMSE scores and prevalences of cognitive impairment in 7,628 participants aged 50 to 89 years at resurvey who reported having had at least some level of formal education, **by district** 3
3. Age- and sex-specific mean MMSE score and prevalence of cognitive impairment in 8,634 participants aged 50 to 89 years at resurvey **including those who reported having no formal education** 4


### Supplementary Figures


1. Prevalence of cognitive impairment and mean MMSE score in 8,634 aged 50 to 89 years at resurvey **including those who reported having no formal education** 5
2. Sex-specific distribution of MMSE scores in 7,628 participants aged 50 to 89 years at resurvey who reported having had at least some level of formal education 6

**Supplementary Table 1. Analysis inclusion criteria**

| | Number of participants |
|---|---|
| **Number of participants who took part in the resurvey** | **10,144** |
| Aged <50 or ≥90 years | 388 |
| MMSE assessment | |
| • Missing MMSE score | 218 |
| • Questionnaire duration <1 or >30 min | 370 |
| Missing data on education | 126 |
| **Remaining participants (sensitivity analysis)** | **9,288** |
| Less those who reported having no formal education | 1,091 |
| **Remaining participants (main analysis)** | **8,197** |

**Supplementary Table 2. Age- and sex-specific mean MMSE scores and prevalences of cognitive impairment in 8,197 participants aged 50 to 89 years at resurvey who reported having had at least some level of formal education, by district**

**Coyoacán**

| | **Men** | | | **Women** | | | **Overall** | | |
|---|---|---|---|---|---|---|---|---|---|
| **Age group** | ***n*** | **MMSE mean (SD)** | **Prevalence %** | ***n*** | **MMSE mean (SD)** | **Prevalence %** | **N** | **MMSE mean (SD)** | **Prevalence %** |
| 50-59 | 353 | 27.2 (2.5) | 11 | 807 | 27.2 (2.7) | 14 | 1160 | 27.2 (2.6) | 12 |
| 60-69 | 457 | 26.7 (3) | 19 | 1116 | 26.4 (3) | 22 | 1573 | 26.6 (3) | 20 |
| 70-79 | 426 | 25.3 (4) | 34 | 654 | 24.8 (3.7) | 38 | 1080 | 25.1 (3.8) | 36 |
| 80-89 | 181 | 22.9 (4.8) | 54 | 254 | 22.3 (4.8) | 63 | 435 | 22.6 (4.8) | 59 |
| **All** | **1417** | **25.5 (3.6)** | **30** | **2831** | **25.2 (3.6)** | **34** | **4248** | **25.4 (3.6)** | **32** |

**Iztapalapa**

| | **Men** | | | **Women** | | | **Overall** | | |
|---|---|---|---|---|---|---|---|---|---|
| **Age group** | ***n*** | **MMSE mean (SD)** | **Prevalence %** | ***n*** | **MMSE mean (SD)** | **Prevalence %** | **N** | **MMSE mean (SD)** | **Prevalence %** |
| 50-59 | 342 | 28 (1.9) | 6 | 1021 | 27.9 (2.3) | 10 | 1363 | 27.9 (2.1) | 8 |
| 60-69 | 347 | 27.3 (2.9) | 12 | 874 | 27 (2.8) | 17 | 1221 | 27.2 (2.8) | 14 |
| 70-79 | 298 | 25.8 (3.7) | 26 | 632 | 25.5 (3.8) | 31 | 930 | 25.6 (3.7) | 28 |
| 80-89 | 161 | 23.7 (4.3) | 49 | 274 | 23 (5.1) | 54 | 435 | 23.3 (4.7) | 52 |
| **All** | **1148** | **26.2 (3.2)** | **23** | **2801** | **25.8 (3.5)** | **28** | **3949** | **26 (3.3)** | **26** |

MMSE, Mini-Mental State Examination. Cognitive impairment defined as MMSE ≤24. Overall prevalences are the simple averages of the age and sex-specific estimates (i.e., are uniformly standardised).

**Supplementary Table 3. Age- and sex-specific mean MMSE score and prevalence of cognitive impairment in 9,288 participants aged 50 to 89 years at resurvey including those who reported having no formal education**

| | Men | | | Women | | | Overall | | |
|---|---|---|---|---|---|---|---|---|---|
| **Age group** | ***n*** | **MMSE mean (SD)** | **Prevalence %** | ***n*** | **MMSE mean (SD)** | **Prevalence %** | **N** | **MMSE mean (SD)** | **Prevalence %** |
| 50-59 | 710 | 27.4 (2.8) | 10 | 1913 | 27.2 (3.1) | 15 | 2623 | 27.3 (2.9) | 13 |
| 60-69 | 846 | 26.6 (3.5) | 19 | 2205 | 26 (3.7) | 26 | 3051 | 26.3 (3.6) | 22 |
| 70-79 | 806 | 25 (4.2) | 35 | 1629 | 23.9 (4.6) | 46 | 2435 | 24.4 (4.4) | 40 |
| 80-89 | 426 | 22.5 (4.8) | 58 | 753 | 20.9 (5.7) | 68 | 1179 | 21.7 (5.3) | 63 |
| **All** | **2788** | **25.4 (3.8)** | **31** | **6500** | **24.5 (4.3)** | **39** | **9288** | **24.9 (4.1)** | **35** |

MMSE, Mini-Mental State Examination. Cognitive impairment defined as MMSE ≤24. Overall prevalences are the simple averages of the age and sex-specific estimates (i.e., are uniformly standardised).

**Supplementary Figure 1. Prevalence of cognitive impairment and mean MMSE score in 9,288 aged 50 to 89 years at resurvey including those who reported having no formal education**

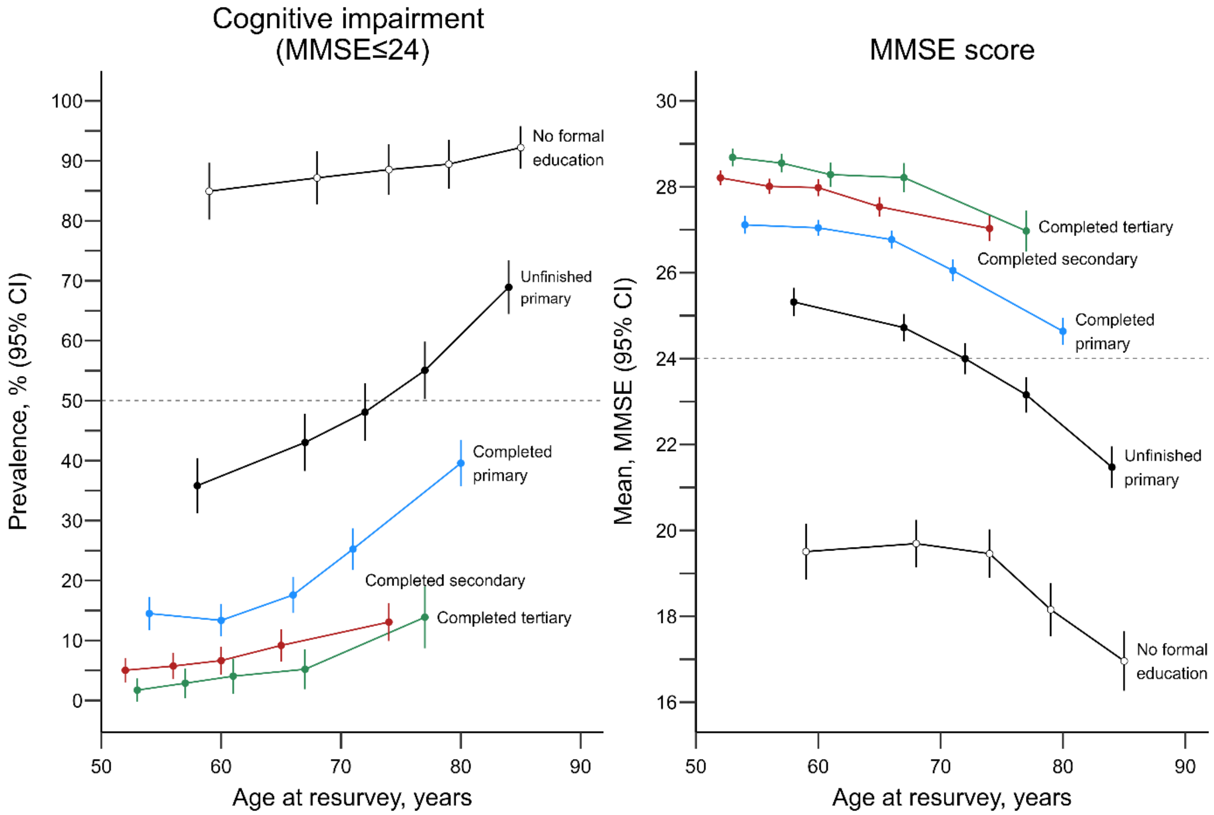


MMSE, Mini-Mental State Examination; CI, confidence interval. Unadjusted prevalences and means with 95% CIs are shown. Each point involves ~210 participants with no formal education, ~410 with unfinished primary, ~610 with completed primary, ~430 with completed secondary, and ~170 with completed tertiary education.

**Supplementary Figure 2. Sex-specific distribution of MMSE scores in 8,197 participants aged 50 to 89 years at resurvey who reported having had at least some level of formal education**

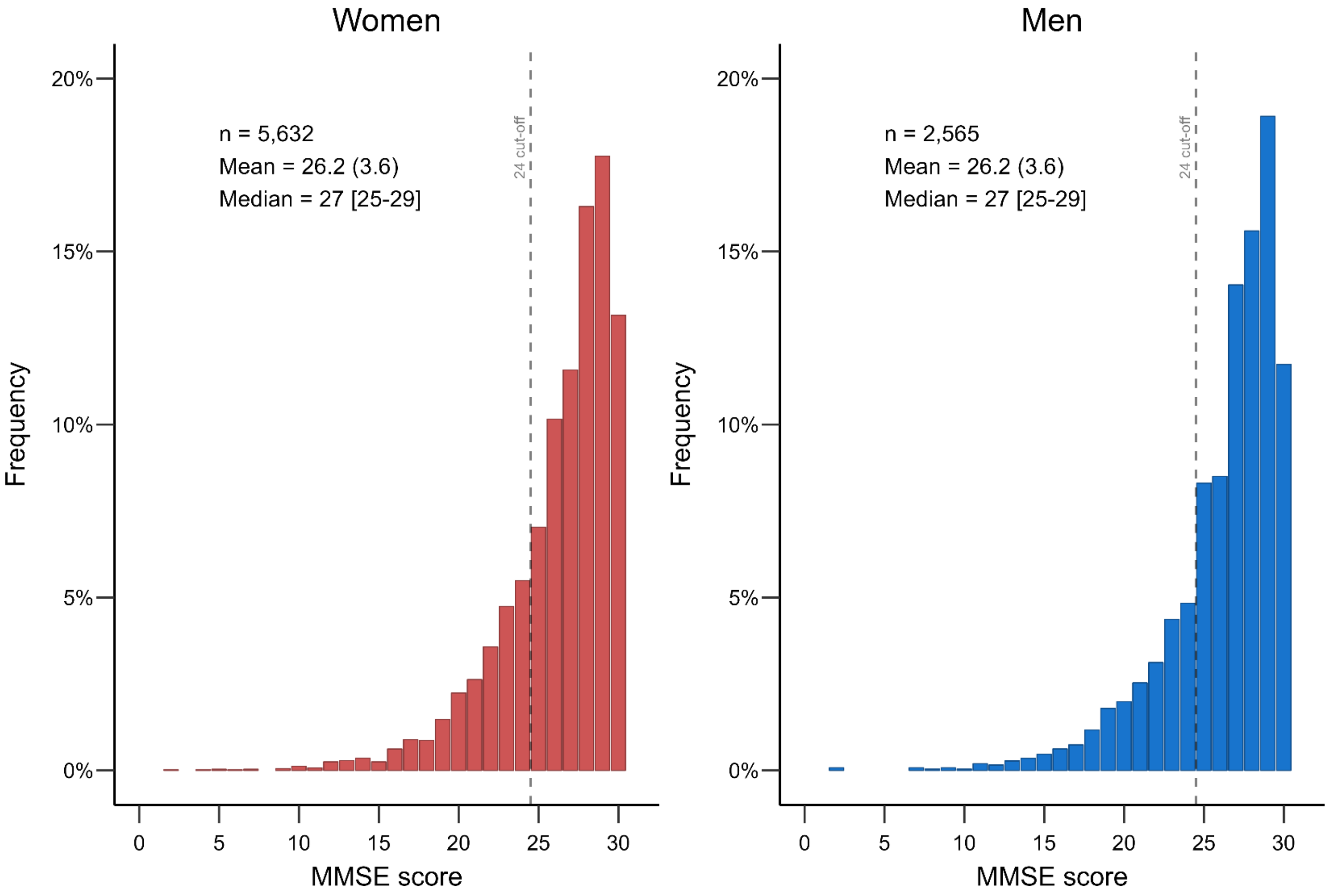


MMSE, Mini-Mental State Examination. Exclusion criteria listed in Supplementary Table 1.
